# Clinical feature, treatment pattern and survival of Esophageal cancer at Tikur Anbessa Specialized Hospital, Ethiopia: a prospective cohort study

**DOI:** 10.1101/2023.02.14.23285932

**Authors:** Jilcha Diribi Feyisa, Adamu Adisse, Eva Johanna Kantelhardt, Girum Tesema Zingeta, Eyoel Negash, Abigiya Wondimagegnewu, Selamawit Hirpha, Muluken Gizaw, Mathewos Aseffa

**Affiliations:** Department of Oncology, Saint Paul Hospital Millennium Medical College, Addis Ababa, Ethiopia; Global Health Working Group, Institute for Medical Epidemiology, Biometrics, and Informatics, Martin-Luther-University Halle-Wittenberg, Halle (Saale), Germany; Department of Oncology, School of Medicine, Addis Ababa University, Addis Ababa, Ethiopia; Department of Preventive Medicine, School of Public Health, Addis Ababa University, Addis Ababa, Ethiopia

**Author notes:** **Corresponding Author** Jilcha Diribi Feyisa, Twitter: @jilchlovmom. **Author Contributions** Jilcha Diribi Feyisa conceptualized the research, was awarded funding for the project, designed the methodology, conducted formal analysis, wrote the original draft and edited subsequent drafts. Mathewos Assefa, Adamu Addissie, and Eva Johanna Kantelhardt were involved in conceptualization, supervision, methodology, and editing. Girum Tesema Zingeta contributed by data collection, data entry and editing. Eyoel Negash contributed by data collection. Selamawit Hirpha, Abigiya Wondimagegnewu and Muluken Gizaw involved in conceptualization, facilitation of funding and ethical clearance for this project.

**Keywords:** Esophageal cancer, clinical feature, treatment pattern, survival, TASH, Ethiopia

## Abstract

**Purpose:** Ethiopia is located within the esophageal belt of Africa. Esophageal cancer is the seventh leading cause of death in Ethiopia. There is a paucity of literature regarding the clinical features, treatment patterns, and survival of patients with esophageal cancer. We report the clinical profile, treatment pattern, and survival of patients with esophageal cancer at Tikur Anbessa Specialized Hospital, Ethiopia.

**Methods:** An unmatched prospective cohort study was conducted from February 27, 2018 to February 28, 2020. We used the Kaplan–Meier method to assess the overall survival time and survival time according to stage and treatment type. Multivariate Cox regression analysis with the backward LR selection method was used to fit the final model.

**Result:** In total, 230 patients with esophageal cancer were recruited for this study. The median survival time was six months (95%CI) (5, 7). A total of 170 (73.9%) patients died during the 1,244 person-month follow-up period, resulting in an overall event rate of 162 per 1,000 person– months. The overall survival rate was very low with 6 months, 1-, and 2-year survival rates of 54.6% (95%CI:47.5%-61.2%), 19.5% (13.8% -25.9%), and 2.0% (0.45%–5.9%), respectively. Chemotherapy, local recurrence, and brain metastases were variables that explained the model.

**Conclusion:** survival of patients with esophageal cancer at Tikur Anbessa Specialized Hospital was very short. We recommend multimodal treatment to improve treatment outcomes.

## Introduction

Ethiopia is an East African country with a population of over 120 million. Esophageal cancer is one of the most common cancers in Ethiopia. In 2020, 1552 new esophageal cases were estimated, and 1478 patients died of the disease in the same year (1). The crude incidence rate of esophageal cancer in the Ethiopian population aged ≥ 15 years per 100,000 people per year was estimated to be 2.4 with an age-specific rate of 3.4 (2).

Based on the seventh edition of the American Joint Committee on Cancer (AJCC) classification for staging, esophageal cancer is categorized into three stages: localized, regional, and distant (3,4). Localized cancer includes stage I and some stage II tumors, and the cancer grows only in the esophagus. Regional means that the cancer has spread to the nearby lymph nodes or tissues. Stages III and IVA are regional diseases. Distant disease include all stage IVB cancers and indicate that the cancer has spread to other organs away from the first tumor (3,4).

Surgery, chemotherapy, radiotherapy, targeted therapy, and endoscopic treatment are the general treatment options for esophageal cancer. The choice of therapeutic modality is primarily dictated by disease stage. Preoperative chemoradiotherapy followed by surgical resection and adjuvant chemotherapy is currently the preferred treatment for resectable tumors without metastasis (cT1-3, No, Mo) (5,6). Primary surgical resection is microscopically positive in approximately 25% of the cases (5). At an advanced stage, resectability decreases, necessitating neoadjuvant and adjuvant chemoradiotherapy (5,7). Surgery alone was associated with significantly better overall survival than other treatment modalities for patients with stage I disease and was associated with significantly worse overall survival in patients with stage III cancer (7). Patients with clinical stage I disease can be treated with esophagectomy, without preoperative therapy. Multimodal treatment is recommended for patients with stage II–III esophageal squamous cell carcinoma (5,7). Neoadjuvant Concurrent chemoradiotherapy before surgery, which is becoming the standard protocol for locally advanced operable esophageal cancer, has better outcomes than surgery alone (5). A pathological complete response of 29 % %was achieved in patients who underwent resection after chemoradiotherapy. The median overall survival was 49.4 months in one study for patients treated with neoadjuvant chemoradiotherapy followed by surgery as compared to 24.0 months only in patients treated with surgery alone (5).

The long-term survival of patients with esophageal cancer, especially for localized disease, has improved over the past few decades (8). In the Taiwanese study group, the 3-year overall survival rates were 60.65% for patients with stage I disease, 36.21% for those with stage II cancer, and 21.39% for patients with stage III carcinoma (22). In China, The five-year survival rate of patients with Esophageal Squamous Cell Carcinoma (ESCC) increased from 3.6% to 21.1% between 1973 and 2010 (8). A study in the United States on the survival of esophageal cancer patients revealed a 10-year survival rate of 14% (9). In Ethiopia, however, the survival of patients with esophageal cancer has not shown any significant improvement over the decades (10). This disease has caused the death of patients in different hospitals, with wide geographical coverage across the nation, which reflects it as a significant health problem.

Literature regarding the clinical features, treatment patterns, and survival of patients with esophageal cancer in Ethiopia is scarce. This study assessed the clinical presentation, treatment pattern, and survival of patients with esophageal cancer at the Tikur Anbessa Specialized Hospital (TASH).

## Materials and Methods

An unmatched prospective cohort study was conducted at TASH in Addis Ababa, Ethiopia, from February 27, 2018 to February 28, 2020. During the first year, patient recruitment was conducted, and follow up and chart evaluation continued into the second year. All eligible, biopsy-confirmed, and newly diagnosed esophageal cancer cases were included. Gastroesophageal junction (GEJ) and/or tumors of the cardia on endoscopy/intraoperative findings, which are biopsy-proven, were included. The site of the lesions, which raises the possibility of gastric cancer compared to esophageal or GEJ cancer, was excluded from the study.

The minimum sample size was calculated using EpiInfo STATCAL, with the following assumptions: a proportion of outcomes (death) of 50% for those who received treatment, 80% power, and a confidence interval of 95%. The total calculated sample size was 206, which was increased by 20% to account for any potential lost charts or incomplete reports, resulting in a sample size of 227. All 230 eligible patients were reviewed during the study period.

Data were collected using a data-abstraction tool. Baseline assessment was conducted at the initial stage and a desk review of relevant documents on the patient’s card was conducted at the TASH. In addition, national health policies, strategies, development plans, clinical care data from health facilities, cancer treatment guides, and other instruments were used to develop the instrument, based on the stated objectives. The tool was standardized in consultation with sub-team members’ experts and re-validated for use in a similar setup. The tool was further pretested on ten patients to ensure that the toll measures what is intended to measure. The tool was slightly modified based on the input obtained from the pre-test.

Two Oncology residents were recruited as the data collectors. Training was provided to the data collectors and one supervisor on the purpose of the study at each phase by the principal investigator to familiarize them with the data collection tool.

Follow up times were conducted at 3, 6, 9, and 12 months. Follow up times were staggered and made according to their regular follow up for their care to minimize participant burden and to ensure that all participants have the opportunity to complete all follow up surveys. At each follow up time, participants were asked to report on a variety of outcomes related to the study, including their symptom status and treatment response.

This study looked at the correlation between dependent and independent variables to determine the treatment outcome of esophageal cancer patients. The dependent variable was the treatment outcome (alive or death), while the independent variables included socio-demographic factors, clinical characteristics, pathologic characteristics, stage at presentation, and treatment types received. These factors were analyzed to determine which variables had the strongest correlation with the patient’s treatment outcome.

Measurements were made for treatment response analysis based on objective measures such as patient-reported symptom improvement, imaging studies, and clinical assessments. Survival analysis was conducted by measuring the length of time from diagnosis to death, or from diagnosis to last follow-up visit. Tumor response was assessed by using Response Evaluation Criteria in Solid Tumors (RECIST criteria). Thus, clinical response to treatment was measured in four ways: complete response (disappearance of all target lesions), partial response (at least a 30% decrease in the sum of the longest diameter of target lesions compared to the baseline), progressive disease (at least a 20% increase in the sum of the longest diameter of target lesions compared to the smallest sum since the treatment started), and stable disease (neither sufficient shrinkage to qualify for partial response nor sufficient increase to qualify for progressive disease compared to the smallest sum since the treatment started).

In order to reduce the risk of bias, careful selection of participants, statistical adjustment of confounding variables, quality control of data collection, regular monitoring of study participants, and regular assessment of outcomes were all employed. Data collection was monitored regularly and double-checked for accuracy, while study participants were monitored regularly and followed-up with if they were at risk of dropping out. Finally, outcomes were assessed and adjusted for any changes in treatment patterns or survival rates. Regular follow-up assessments and contact attempts were conducted with participants to address loss to follow-up. Mobile phone contact was made at regular intervals, and the baseline characteristics of participants were compared to ensure no major discrepancies between those followed up and those lost to follow-up.

After checking the data for completeness and consistency, they were entered into the Epi Data Manager version 4.6.0.0. The entered data were cleaned and exported to SPSS, version 25, for analysis. STATA Version 14 was used for Kaplan-Meier survival analysis. Descriptive statistics was used to summarize the data collected at each follow up time to compare the differences in outcomes between the baseline and follow up periods. Additionally, Kaplan-Meier analysis was used to determine the survival of patients with esophageal cancer. Bivariate Cox regression analysis was performed to identify the possible predictors of survival in patients with esophageal cancer. Multivariate Cox regression analysis was used to determine the net effect of each predictive factor, after controlling for possible confounding factors. Variables with a log rank value < 0.05 on Kaplan Meir and the omnibus tests of model < 0.05 on bivariate Cox regression were considered to have a significant association. A multivariate Cox proportional hazard regression model was used to control for confounders and assess the survival of patients with esophageal cancer. The assumption of proportional hazard was checked using the Shenfield residual proportional hazard test, the presence of multicollinearity was checked, and then a model adequacy test was performed to test the goodness of fit. Missing data were addressed by using multiple imputation (MI) techniques. Furthermore, sensitivity analyses were conducted by varying assumptions and the definition of the exposure or outcome to assess the robustness of the results to the assumptions of MI.

Ethical clearance was obtained from the Addis Ababa University Institutional Review Board (IRB) (IRB Reference Number: 096/17/SPH). Permission to review patient charts and contact patients during their visits was obtained from the oncology department. Written informed consent to participate in the study was obtained during the hospital visits. Confidentiality of the information was maintained throughout the study by excluding names as identification in the data extraction form and the data used only for the purpose of the conducted study. Authors did not have access to any information that could identify individual participants at any stage of the data collection process.

## Result

### Sociodemographic Characteristics and Geographic distribution

A total of 230 esophageal cancer patients diagnosed and treated between January 1, 2018, and January 1, 2020, were recruited for the study. Of those, 36 completed all four follow up visits over 12 months, 98 completed three follow up visits over 9 months, 102 completed two follow up visits over 6 months, and 183 presented for the first follow up over 3 months. All 230 individuals have been analyzed for study. A phone assessment was conducted for 187 patients at the end of their follow up to assess their final status. Forty-three patients were unable to receive a phone assessment due to the difficulty of assessing patients or care givers over the phone.

Of 230 patients recruited, 130 (56.5%) were females and 100 (43.5%) were males. The female-to-male ratio was 1.3:1. The mean age of the participants was 52 (SD = 13) years. Their ages ranged from 22 to 85 years old. The highest incidence of esophageal cancer occurred in 148 (64.3%) patients occurred between age range of 46-70 years. More than half of the patients 121 (52.6%) were Muslim, followed by Orthodox Christianity 81 (35.2%). The majority of 189 (82.2%) patients were married. With respect to occupation, 100 (43.5%) were farmers, and 92 (40%) were housewives (Table 1).

**Table 1:**
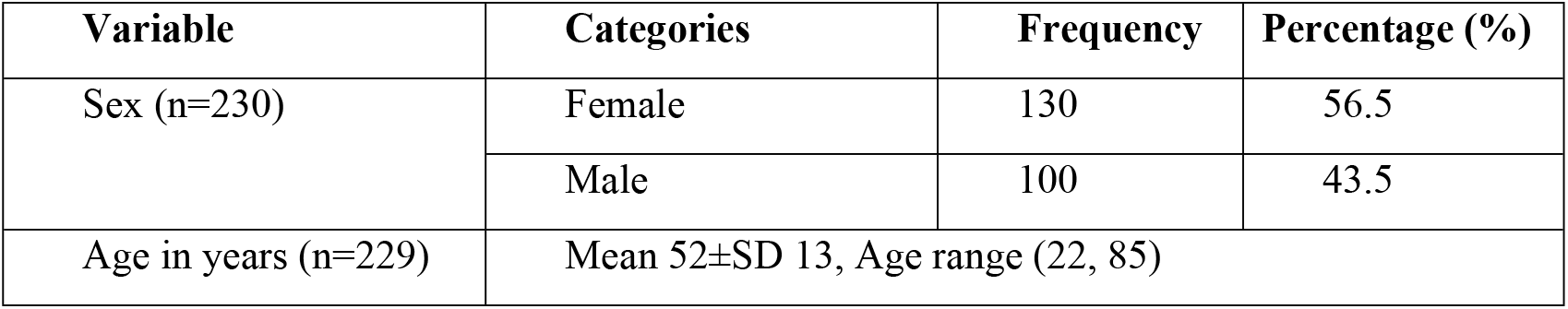
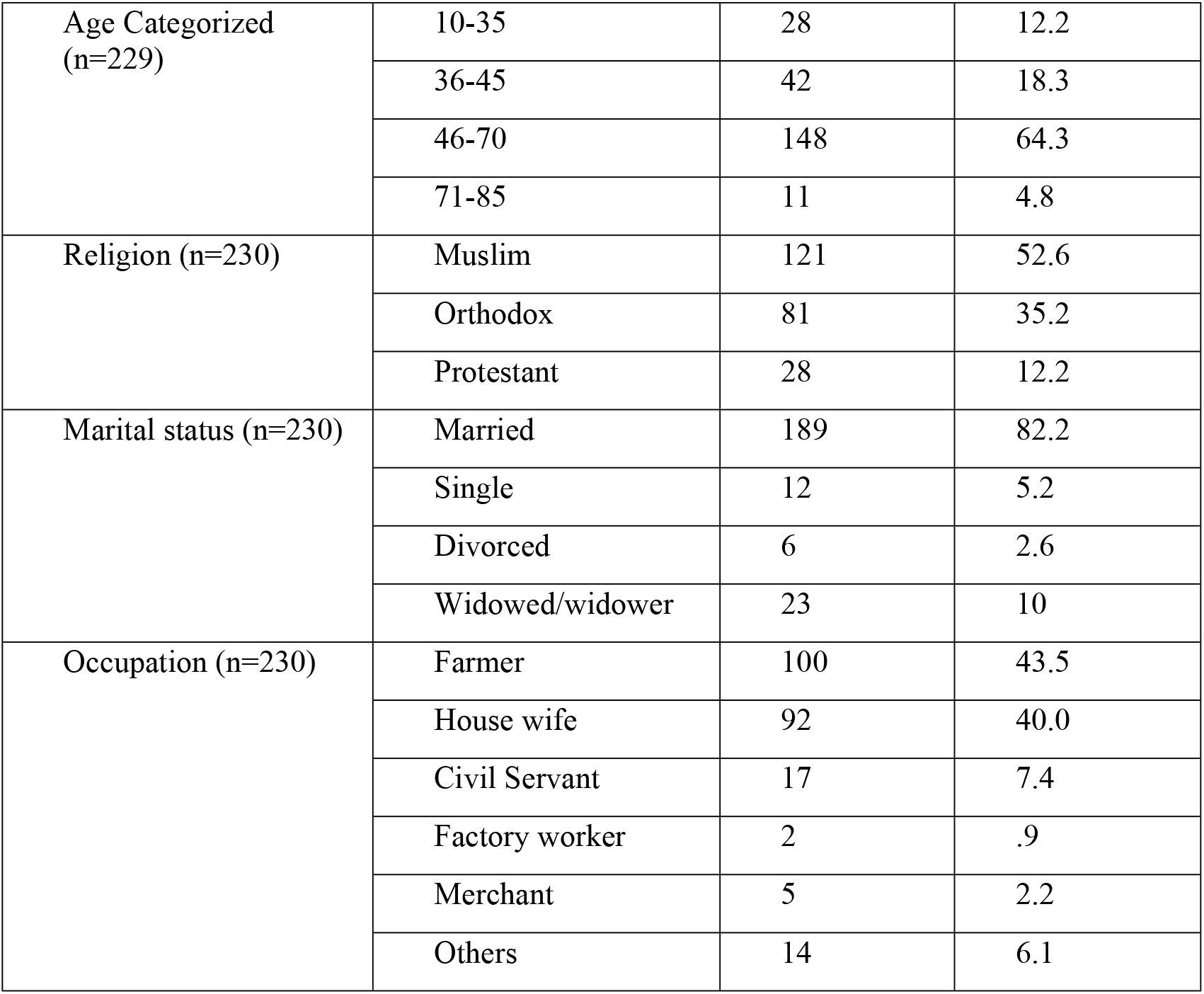
Sociodemographic of esophageal cancer patients at Tikur Anbessa Specialized Hospital, Addis Ababa, Ethiopia 2018 – 2020

| Variable | Categories | Frequency | Percentage (%) |
| --- | --- | --- | --- |
| Sex (n=230) | Female | 130 | 56.5 |
|  | Male | 100 | 43.5 |
| Age in years (n=229) | Mean 52±SD 13, Age range (22, 85) |  |  |
| Age Categorized<br>(n=229) | 10-35 | 28 | 12.2 |
|  | 36-45 | 42 | 18.3 |
|  | 46-70 | 148 | 64.3 |
|  | 71-85 | 11 | 4.8 |
| Religion (n=230) | Muslim | 121 | 52.6 |
|  | Orthodox | 81 | 35.2 |
|  | Protestant | 28 | 12.2 |
| Marital status (n=230) | Married | 189 | 82.2 |
|  | Single | 12 | 5.2 |
|  | Divorced | 6 | 2.6 |
|  | Widowed/widower | 23 | 10 |
| Occupation (n=230) | Farmer | 100 | 43.5 |
|  | House wife | 92 | 40.0 |
|  | Civil Servant | 17 | 7.4 |
|  | Factory worker | 2 | .9 |
|  | Merchant | 5 | 2.2 |
|  | Others | 14 | 6.1 |

Dysphagia was the most common 146 (63.5%) first symptom observed by the patient followed by heart burn 46 (20%) and epigastric burning pain 18 (7.8%). Almost all patients (n = 229, 99.6%) had dysphagia during presentation. The other common symptoms associated with esophageal cancer presentation were weight loss 223 (97%), heart burn 175 (76.1%), epigastric burning pain 143 (62.2%), vomiting 136 (59.1%), cough 19 (8.3%), chest pain 13 (5.7%), and hoarseness of voice 2 (0.9%). The mean amount of weight lost during presentation was 10.3Kg ± SD 6.8 kg. Of 229 patients evaluated during the study period, 134 (58.3%) presented with performance status of Eastern Cooperative Oncology Group (ECOG) I followed by 76 (33%) with ECOG II. Seventeen (7.4%) patients presented with a poor performance status (ECOG III). Of 230 patients, 185 (80.4%) of the study participants had squamous cell carcinoma (SCC) and 30 (13%) had adenocarcinoma (AC) type of esophageal cancer. Of the 14 patients who underwent esophagectomy, 7 (50%) had involvement of the muscularis propria, 6 (2.6%) had involvement of the adventitia, and 1 (7.1%) had involvement of the submucosa. A free surgical margin was achieved in 9 (64.3%) cases. Three (21.5%) cases had positive margins, while margin status was not reported in two (14.3) cases (Table 2).

**Table 2:** Clinical and pathologic feature of esophageal cancer patients at Tikur Anbessa Specialized Hospital, Addis Ababa, Ethiopia 2018 – 2020

| **Variable** | **Categories** | **Frequenc**<br>**y** | **Percentage**<br>**(%)** |
| --- | --- | --- | --- |
| Comorbid Illness<br>(n=229) | No | 212 | 92.2 |
|  | RVI | 7 | 3.0 |
|  | HTN | 6 | 2.6 |
|  | DM | 2 | 0.9 |
|  | Others | 3 | 1.3 |
| First observed<br>symptom (n=229) | Dysphagia | 146 | 63.5 |
|  | Heart burn | 46 | 20.0 |
|  | Epigastric pain | 18 | 7.8 |
|  | Vomiting | 15 | 6.5 |
|  | Weight Loss | 3 | 1.3 |
|  | Chest Pain | 1 | 0.4 |
| Presenting Symptoms<br>(n=229) | Dysphagia | 229 | 99.6 |
|  | Weight loss | 223 | 97 |
|  | Heart burn | 175 | 76.1 |
|  | Epigastric burning pain | 143 | 62.2 |
|  | Vomiting | 136 | 59.1 |
|  | Cough | 19 | 8.3 |
|  | chest pain | 13 | 5.7 |
|  | Hoarseness of voice | 2 | 0.9 |
| Performance Status<br>ECOG (229) | 0 | 2 | 0.9 |
|  | I | 134 | 58.3 |
|  | II | 76 | 33.0 |
|  | III | 17 | 7.4 |
| Biopsy (n=229) | Yes | 226 | 98.3 |
|  | No | 3 | 1.3 |
| Type of Biopsy<br>(n=224) | Endoscopic | 210 | 93.8 |
|  | Esophagectomy | 14 | 6.3 |
| Histologic subtype<br>(n=226) | SCC | 185 | 80.4 |
|  | AC | 30 | 13.0 |
|  | ASC | 1 | 0.4 |
|  | Carcinoma, NOS | 10 | 4.3 |
| Histologic Grade<br>(n=226) | Not reported | 186 | 80.9 |
|  | Gx | 8 | 3.5 |
|  | G1 | 17 | 7.4 |
|  | G2 | 3 | 1.3 |
|  | G3 | 12 | 5.2 |
| Esophageal layer<br>involved - for those<br>esophagectomy was<br>done (n=14) | Submucosa | 1 | 7.1 |
|  | Muscularis propria | 7 | 50 |
|  | Adventitia | 6 | 42.9 |
| Margin status - for<br>those esophagectomy<br>was done (n=14) | Free | 9 | 64.3 |
|  | Involved | 3 | 21.5 |
|  | Not reported | 2 | 14.3 |

### Stage at presentation

The clinical stage at presentation was recorded in 196 and 25 patients with SCC and AC, respectively. Of the 196 patients with SCC, 93 (47.4%) had staged IVA (Fig 1). Metastases (stage IVB) were diagnosed in 68 patients (34.7%). The remaining patients were diagnosed as stage III (n = 19, 9.7%) and stage II (n = 14, 7.1%). Only 2 (1%) patients presented with stage I disease. Fifteen (60%) out of the 25 patients with AC presented with metastatic disease (stage IVB). Six (24%) patients were diagnosed with stage IVA, two (8%) patients were diagnosed with stage III, and two (8%) were diagnosed with stage I. Fourteen patients underwent esophagectomy; hence, pathologic staging was performed only for 14 patients. Of the 14 patients, six (42.9%) presented at T2 and six (42.9%) presented at T3. Two patients had T1b and T4a tumors on pathological evaluation. Of the 14 patients who underwent esophagectomy, 13 had SCC. Pathological group staging of patients with SCC revealed stages IB (7.7%), IIA (23.1%), IIB (7.7%), IIIA (15.4%), IIIB (38.5), and IVA (7.7%). One patient with AC stage IIIB AC disease (Fig 1). (For details on the stage at presentation, see Supplemental table 1).

**Fig 1:**
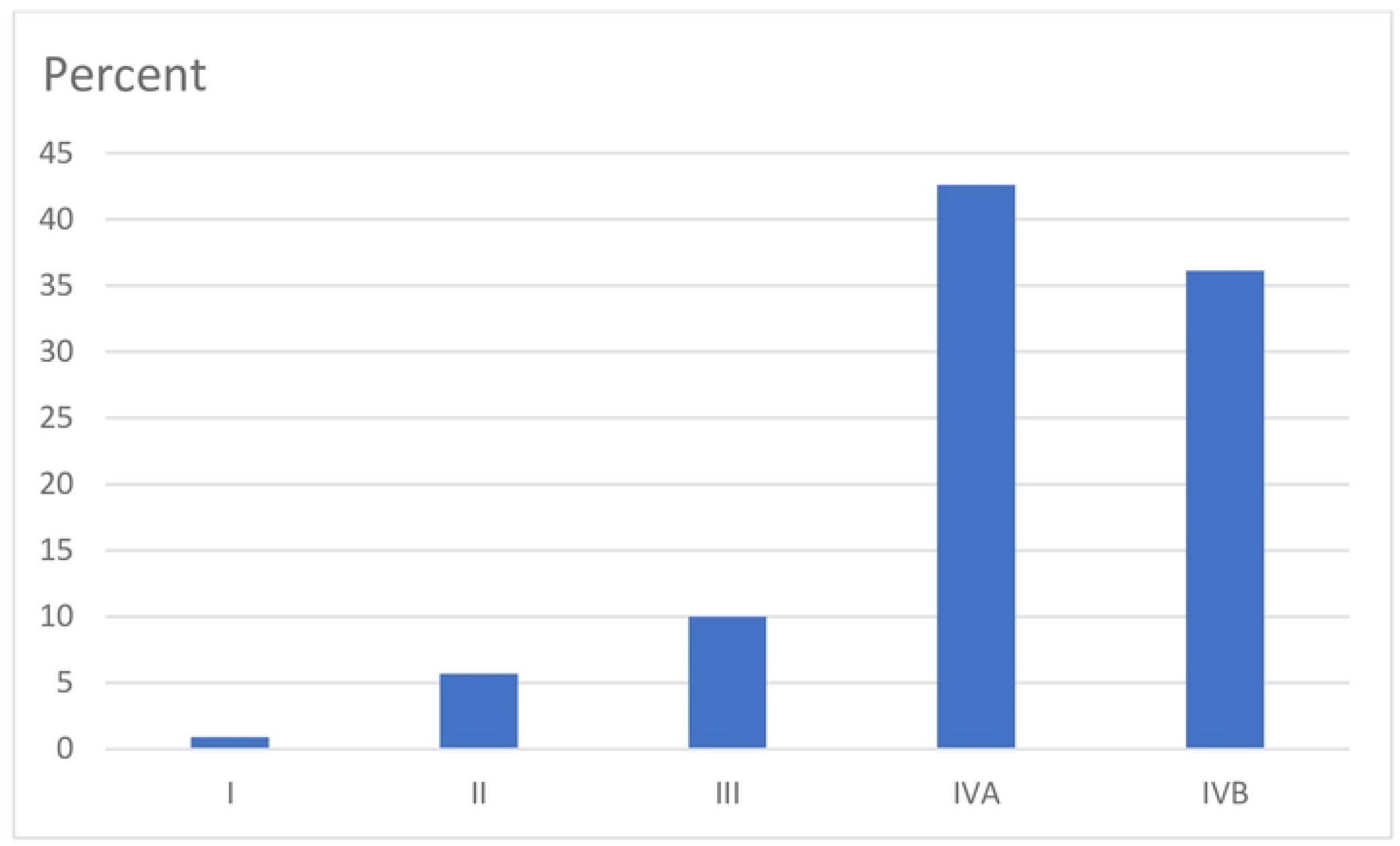
Stage at presentation of esophageal cancer patients at Tikur Anbessa Specialized Hospital, Addis Ababa, Ethiopia 2018 – 2020.

### Treatment Pattern

Regarding the type of treatment received, 68 (70.1%) were prescribed chemotherapy, followed by palliative radiotherapy (n=15, 15.6%) and surgery (n=14, 14.4%) patients (Fig 2). A feeding tube was inserted in 42 patients (18.3%). Of the 15 patients who received radiotherapy, only one (6.7%) received definitive radiotherapy, whereas the remaining 14 (93.3%) received radiotherapy for palliation of symptoms. Most patients 170 (74.2%) did not receive any kind of surgical intervention. The intent of chemotherapy was palliative in 60 patients (69.3%). Three (4.5%) patients received neoadjuvant chemotherapy, two (3%) received adjuvant chemotherapy after esophagectomy, and only one (1.5%) received concurrent chemotherapy with radiotherapy. Cisplatin plus 5 fluorouracil was the most common chemotherapy regimen received (n=43, 70.2%), followed by cisplatin plus paclitaxel (n=22, 32.8%). Of the 67 patients who received chemotherapy, one-third (34.3%) completed 6 cycles of chemotherapy. Fifteen (22.4%) patients received three cycles of chemotherapy and 13 (19.4%) received only one cycle of chemotherapy. The intent of radiotherapy was palliative in the majority of cases 12 (80%). Only two patients (13.3%) received neoadjuvant chemotherapy with the intent of operating after chemotherapy. Of the 15 (6.5%) patients who received radiotherapy, only one (6.7%) received a radical dose (66 Gy in 33 fractions). The other 1 (6.7%) received 40 Gy in 20 fractions concurrently with chemotherapy. All remaining patients received radiotherapy for palliation. Trans-hiatal esophagectomy was the most common surgical approach used for esophagectomy (n = 9, 64.3 %), followed by Mk Kewon Esophagectomy (n = 4, 28.6%). Ivor Lewis Esophagectomy was performed in one patient (7.1%). Out of 14 patients who underwent esophagectomy, a free surgical margin was achieved in 9 (64.3%). Three (21.5%) cases had positive margins, while margin status was not reported in two (14.3) cases. (For details of the treatment received, see Supplemental table 2).

**Fig 2:**
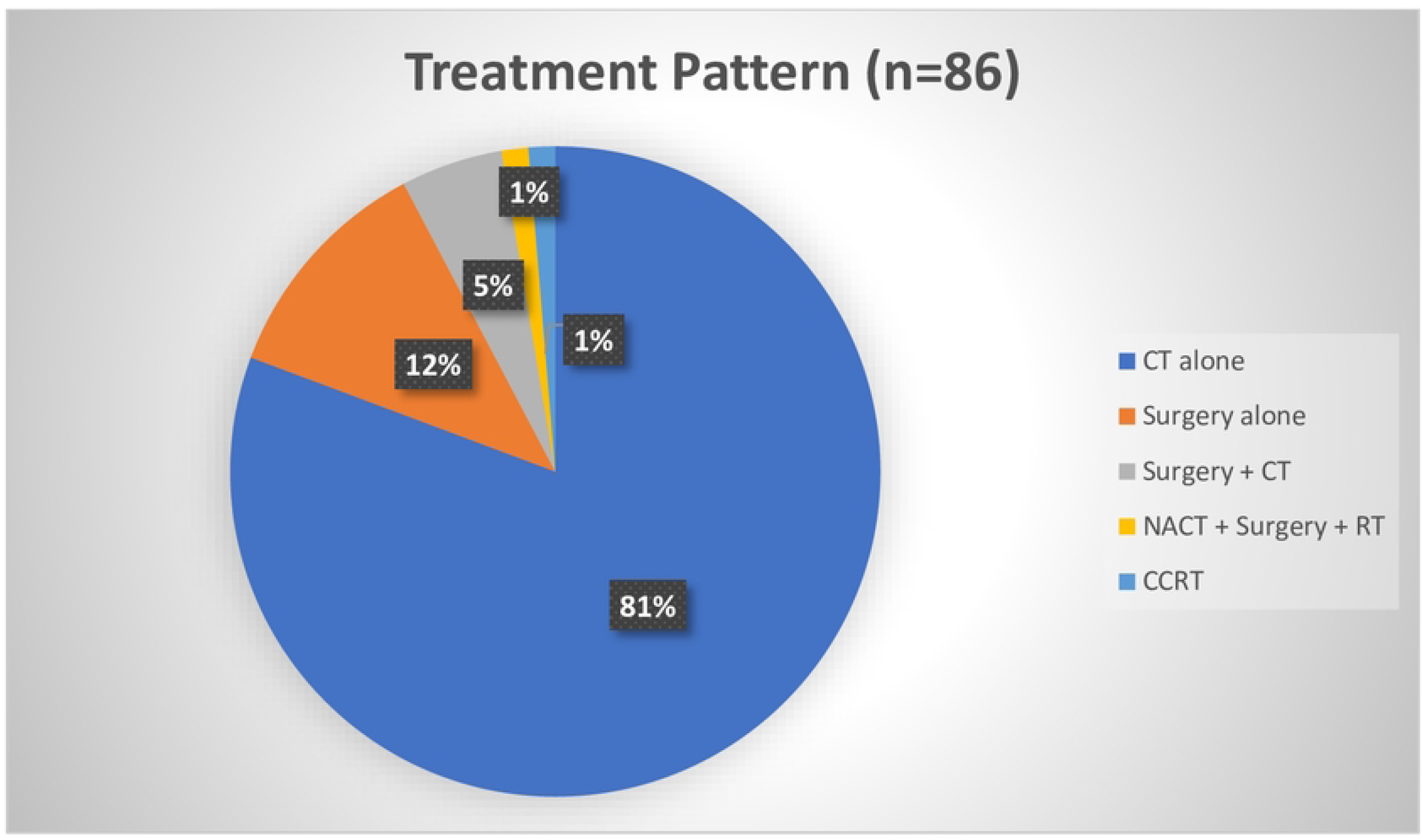
Treatment pattern for esophageal cancer patients at Tikur Anbessa Specialized Hospital, Addis Ababa, Ethiopia 2018 – 2020

### Outcome

Nine patients (3.9%) developed distant metastasis during the follow-up period. The lung was the most common site of metastases 6 (66.7%), followed by the liver 2 (22.2%) and vertebrae 1 (11.1%). The occurrence of metastases was unknown in 180 (78.3%) patients. Eight patients (3.5%) developed local recurrence. Six (2.6%) patients had a partial response to treatment, 15 (6.5%) were in stable condition, and 24 (10.4%) had progressive disease. Treatment response was not assessed in 183 (79.6%) patients.

### Overall survival from time of diagnosis to death

The mean and median survival times were 7 ± SD (0.5) months 95%CI (6, 8) and 6 months 95%CI (5, 7), respectively. The overall survival rate showed a decline starting from the early months, as shown by the Kaplan-Meier curve and time table (Fig 3, Table 3). The median follow-up duration was 35 months (IQR, 15–42 months). A total of 170 (73.9%) patients died during the 1,244 person-month follow-up period, resulting in an overall event rate of 162 per 1,000 person–months. The overall survival rate was very low, with six months, one-, and two-year survival rates of 54.6% (95%CI:47.5%-61.2%), 19.5% (13.8% -25.9%), and 2.0% (0.45%– 5.9%), respectively. Only 17 (7.4%) patients were alive at the time of analysis. The status of 43 (18.7%) patients was unknown. Of the 170 patients who died, 166 (72.2%) died at home, while the remaining four (1.7%) died in the hospital.

**Table 3:** Survival time table of esophageal cancer patients at Tikur Anbessa Specialized Hospital, Addis Ababa, Ethiopia 2018 – 2020

| Interval | Beg. | Deaths | Lost | Survival | Std. | 95%CI |
| --- | --- | --- | --- | --- | --- | --- |

|  |  | **Total** |  |  |  | **Error** |  |  |
| --- | --- | --- | --- | --- | --- | --- | --- | --- |
| 0 | 6 | 222 | 91 | 43 | **54.61** | 3.52 | 47.46 | 61.20 |
| 6 | 12 | 88 | 55 | 5 | **19.48** | 3.09 | 13.83 | 25.87 |
| 12 | 18 | 28 | 21 | 3 | **4.04** | 1.66 | 1.62 | 8.25 |
| 18 | 24 | 4 | 2 | 0 | **2.02** | 1.31 | 0.45 | 5.97 |
| 24 | 30 | 2 | 1 | 0 | **1.01** | 0.97 | 0.10 | 4.73 |
| 30 | 36 | 1 | 0 | 1 | **1.01** | 0.97 | 0.10 | 4.73 |

**Fig 3:**
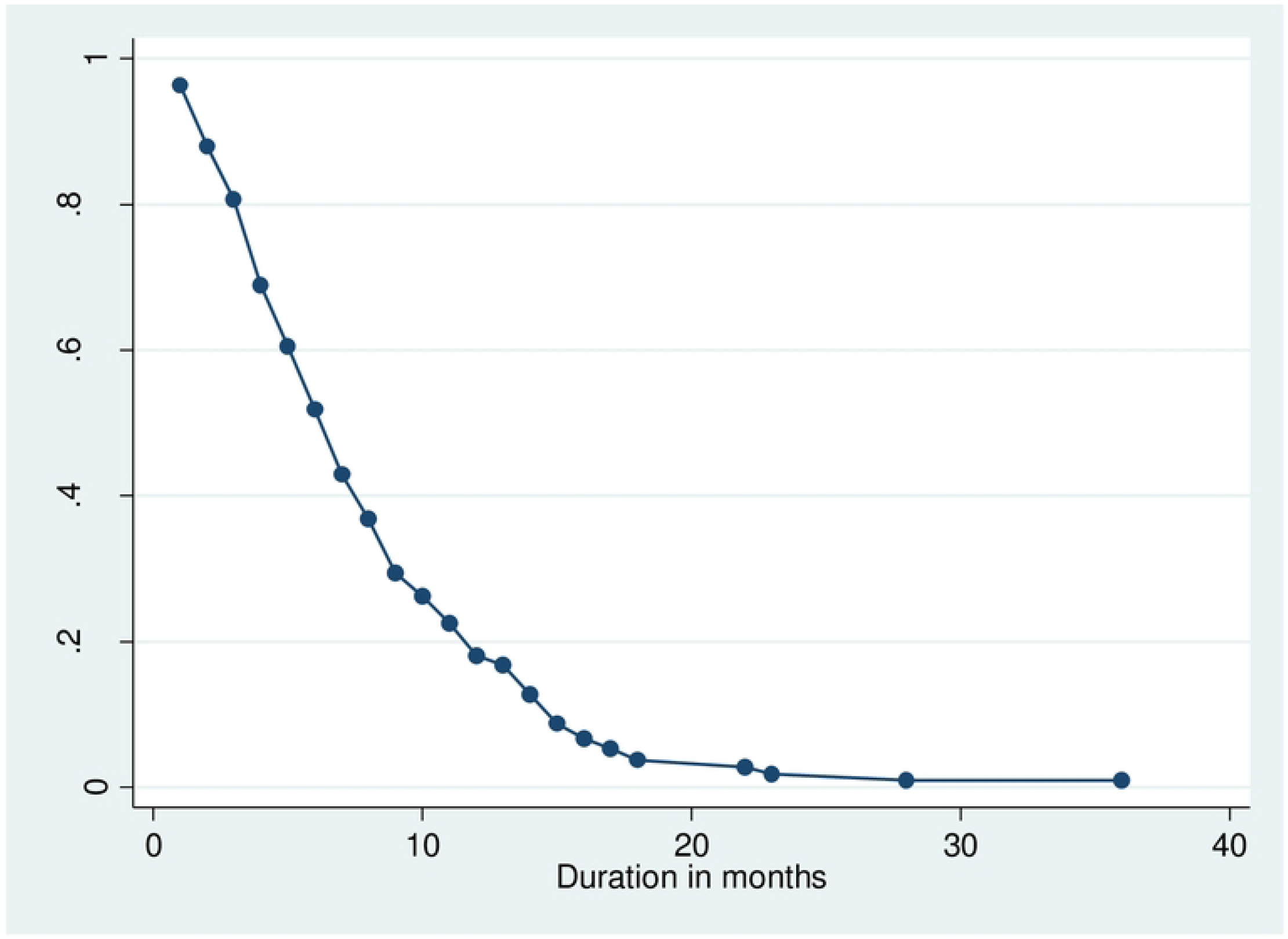
Kaplan Meier plot of overall survival function of esophageal cancer patients at Tikur Anbessa Specialized Hospital, Addis Ababa, Ethiopia 2018 – 2020. The curve shows the median survival is 7 months.

### Survival experience among different groups of esophageal cancer patients

The survival time varied among different categories of covariates, such as pathological and clinical stage at presentation (see Supplemental Fig) and treatment with surgery, chemotherapy, and/or radiotherapy (Fig 4 A-C). The median survival times for stages I, II, III, IVA, and IVB were 8, 4 (95%CI:2-5.7), 9 (6.6 -11), 6 (4.8-7), and 4 (2.6–5.4) respectively. Patients with no distant metastases (IVA) during diagnosis had better survival than those with distant metastases (IVB) (log-rank test, P = 0.0158) (see Supplemental Fig).

**Fig 4(A-C):**
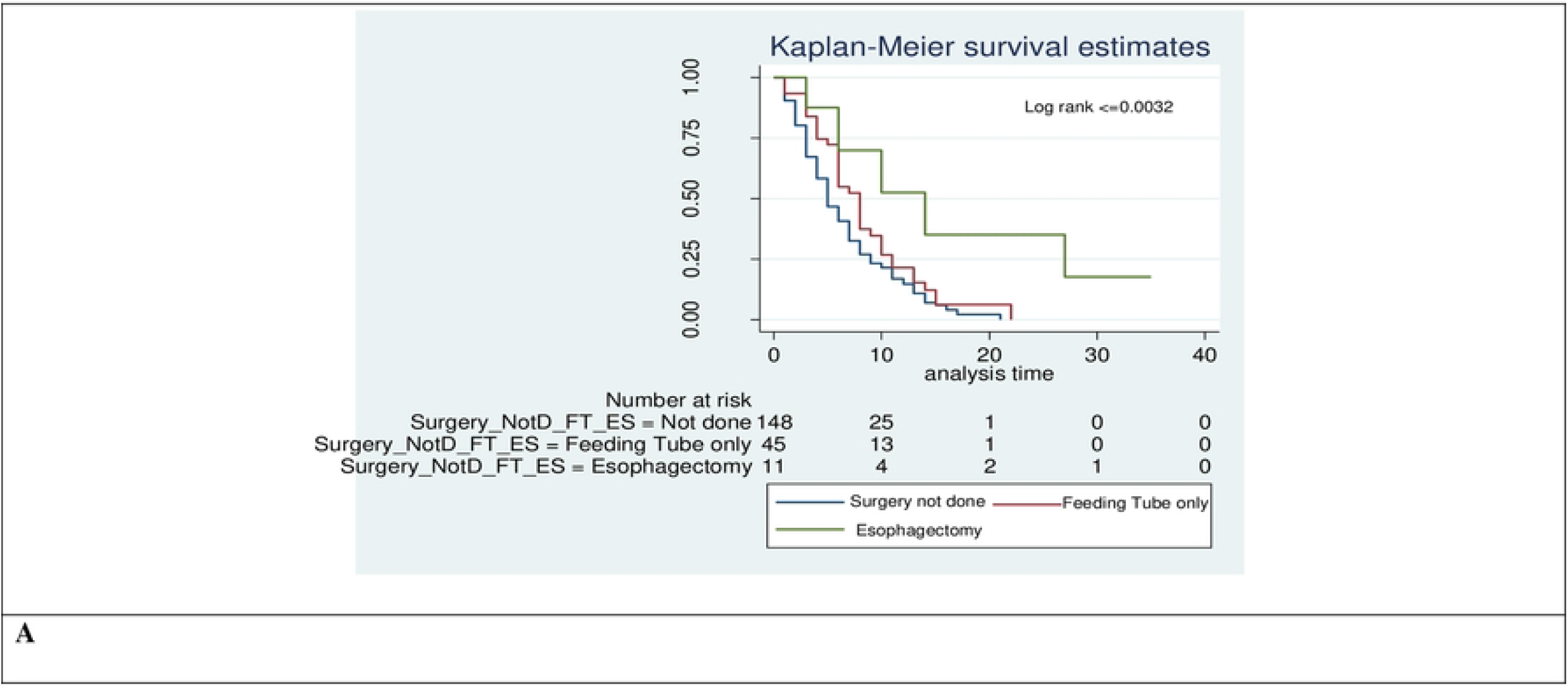
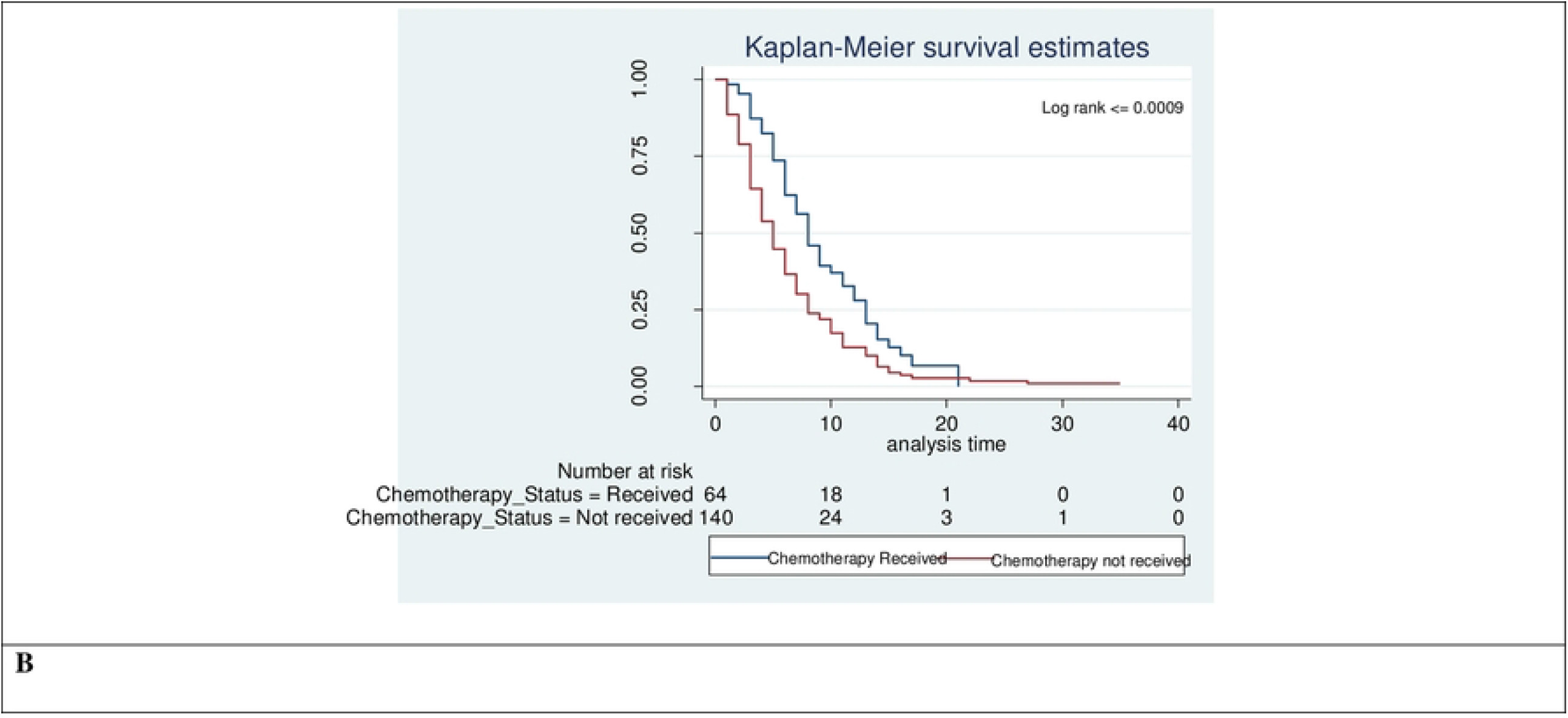
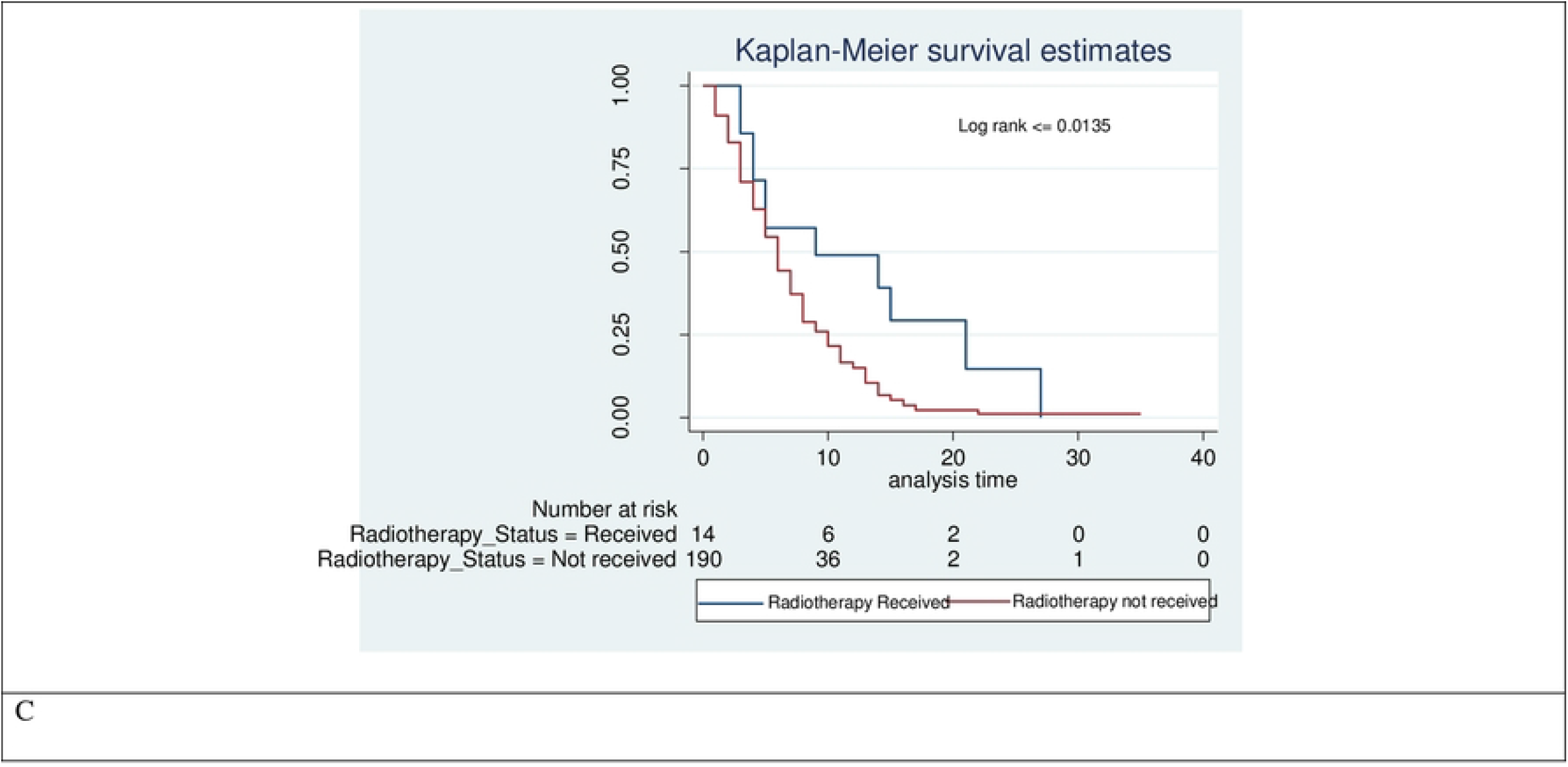
Kaplan Meier plot of survival experience of esophageal cancer patients based on treatment received at Tikur Anbessa Specialized Hospital, Addis Ababa, Ethiopia 2018 – 2020.

Patients who underwent esophagectomy had better survival rates than those who did not (log-rank test, P = 0.0032) (Fig 4 A). Treatment with chemotherapy and/or radiotherapy was associated with improved survival in Kaplan-Meier analysis (log-rank, P= 0.009 and 0.0135, respectively) (Fig 4 B and C).

### Bivariate association between survival and independent variables

On bi-variable analysis, performance status, atrial involvement, distant metastases, brain metastases, clinical stage SCC, treatment with chemotherapy, cycles of chemotherapy given, sequence of treatment given, occurrence of metastases during follow-up, local recurrence, and clinical response after treatment were significantly associated with survival of esophageal cancer patients and selected for multivariate Cox regression with a value <0.05 (see Supplemental Table 3).

### Multicollinearity test

A multicollinearity test was performed, and multicollinearity was suspected for two variables: the sequence of treatment administered (T=-1.945) and metastases during follow-up (T=-2.214, VIF 34.993). These two variables were removed from the list and the other variables were included in the final model (see Supplemental Table 4).

### Multivariable association between survival and independent variables

The final model for patients with esophageal cancer was established for 170 patients. The final model was significant (X_2_ (10) = 37.331, P=0.000). Performance status at presentation, treatment with chemotherapy, local recurrence, and brain metastases were the variables that explained the model. Patients who didn’t received chemotherapy were 7.25 times more likely to die than patients who received chemotherapy, with AHR and 95% CI 7.25, 95% CI 2.57 – 20.48). Patients with no local recurrence were more likely to survive patients with local recurrence with AHR 0.56% CI 0.33 – 0.96. Patients with brain metastases were 14.96 times more likely to die than patients with no brain metastases, with AHR 14.96, 95% CI 1.27 – 174.2 (see supplemental Table 5).

## Discussion

A total of 230 esophageal cancer patients diagnosed between February 27, 2018, and February 28, 2020, were recruited for the study. After initial recruitment, 183 patients presented for the first follow up, 102 for the second follow up, 98 for the third follow up, and only 36 for the fourth follow up at 3, 6, 9, and 12 months. This significant drop in attendance at each follow up was mainly due to clinical deterioration or death. Additionally, some patients chose to receive palliative treatment at their local primary hospitals due to their health condition and financial or transportation-related concerns.

The mean age of the participants was 52 (SD = 13) years. This is lower than the age reported in Southeast Asia and Western countries but almost similar to the mean age (57.8+_11.7SD) reported in Ghana (7). More than half of the patients 121 (52.6%]) were Muslims. This is inconsistent with previous studies that showed that cancer of the oral structures, pharynx, larynx, and esophagus is also generally quite low in religious societies (11).

In agreement with other studies, dysphagia was the most common 146 (63.5%) first symptom observed by the patient (12). Almost all patients (n = 229, 99.6%) had dysphagia during presentation. Heartburn (n = 46, 20%) and epigastric burning pain (n = 18, 7.8%) were the two most common first presenting symptoms. This is a striking finding, as it has not been reported previously. This will have implications for heartburn and epigastric burning pain, enabling early screening of the disease.

More than two third, 185 (80.4%) of the study participants had squamous cell carcinoma (SCC) and 30 (13%) had adenocarcinoma (AC) type of esophageal cancer. This is in line with previous findings that have established a high incidence of ESCC in Africa and Asia (13). An Ecological Study of the African ESCC corridor - easterly African countries stretching from Sudan south to the Eastern Cape Province of South Africa–revealed similar results (14).

The clinical stage at presentation was recorded in 196 and 25 patients with SCC and AC, respectively. Similar to most studies in other low-income countries, most patients presented with advanced disease belonging to either stage III or IV (15). A total of 119 patients had distant metastases to the liver 44 (19.1%), lung 34 (14.8%), celiac lymph nodes 24 (10.4%), peritoneum 7 (3%), supraclavicular lymph node 4 (1.7%), vertebrae or bone 5 (2.2%) or brain 1 (0.4%). Of the 196 patients with SCC, 93 (47.4%) had staged IVA. Metastases (stage IVB) were diagnosed in 68 patients (34.7%). The remaining patients were diagnosed as stage III (n = 19, 9.7%) and stage II (n = 14, 7.1%).

Definitive surgery, neoadjuvant chemoradiotherapy followed by surgery and adjuvant chemotherapy, and concurrent chemoradiotherapy are recommended options for the treatment of esophageal cancer, depending on the stage at presentation and the site of the lesion (5,16). Combined therapy has been established as the preferred treatment modality after significantly increasing overall survival compared to a single modality (5,17–20). Survival rates at one year and five years were better among patients receiving combined treatment than among those undergoing surgery (21). Chemoradiotherapy followed by surgery has been reported in many studies to be associated with better survival and quality of life (22). In the present study, chemotherapy was commonly prescribed to 68 (70.1%) patients, followed by palliative radiotherapy 15 (15.6%) and surgery 14 (14.4%). In contrast to the recommendation, almost all patients who received treatment received monotherapy rather than the recommended multimodal treatment (22).

Sixty-seven patients underwent chemotherapy. The intent of chemotherapy was palliative in 60 patients (69.3%). Three (4.5%) patients received neoadjuvant chemotherapy, two (3%) received adjuvant chemotherapy after esophagectomy, and only one (1.5%) received concurrent chemotherapy with radiotherapy. Cisplatin with 5 fluorouracil was the most common 43 (70.2%), followed by cisplatin with paclitaxel (n = 22, 32.8%). Similar chemotherapy regimens have been reported previously (23).

Fifteen patients were treated with RT. Of the 15 patients who received radiotherapy, only one (0.4%) received a definitive dose (66 Gy) of radiotherapy. Fourteen patients underwent an esophagectomy. Most patients 170 (74.2%) did not receive any kind of surgical intervention. This is very low compared with that in other countries. Among esophageal cancer patients treated in Korea, surgery alone was performed in 31.3%, followed by definitive concurrent chemoradiotherapy (CCRT) (27.0%), neoadjuvant therapy (12.4%), adjuvant therapy (11.1%), and endoscopic resection (5.8%) (20). Trans hiatal esophagectomy was the most common surgical approach used for esophagectomy (n = 9, 64.3 %), followed by Mk Kewon Esophagectomy (n = 4, 28.6%). Ivor Lewis Esophagectomy was performed in one patient (7.1%). This is contrary to the choices reported for other surgical approaches. Extended transthoracic esophagectomy achieved a higher rate of R0 (complete) resection, higher lymph node yield, and prolonged survival than trans hiatal esophagectomy in pT3, cT3, and node-positive patients (41). Out of 14 patients who underwent esophagectomy, a free surgical margin was achieved in 9 (64.3%). Three (21.5%) cases had positive margins, while margin status was not reported in two (14.3) cases.

In this study 8 (3.5%) developed local recurrence during follow-up. However, this is not representative because only 14 patients underwent esophagectomy, and most of them were not followed up. Both locoregional recurrence and hematogenous metastases frequently develop in esophageal cancer even after complete resection (24).

Nine patients (3.9%) developed distant metastasis during the follow-up period. The lung was the most common site of metastases 6 (66.7%), followed by the liver 2 (22.2%), vertebrae 1 (11.1%) and brain 1 (11.1%). This is consistent with autopsy findings; the lung (31%), liver (23%), and bone (13%) were also the most common sites, and brain metastasis was still rare (less than 5%) (43). However, this is different from two other studies that reported the liver as the most common metastatic site in patients with esophageal cancer, followed by the lungs, bones, and the brain (25,26). The occurrence of metastases was unknown in 180 (78.3%) patients. This is because patients succumb to the disease early or are lost to follow-up, making it difficult to assess metastatic status.

Of the 230 patients, 170 (73.9%) died during the 1,244 person-month follow-up period, resulting in an overall event rate of 162 per 1,000 person–months. The mean and median survival times were 7 ± SD (0.5) months 95%CI (6, 8) and 6 months 95%CI (5, 7), respectively, which were lower than the median survival of patients with esophageal cancer reported in the literature (5,9). The overall survival rate declined in the early months. The overall survival rate was very low, with six months, one-, and two-year survival rates of 54.6% (95%CI:47.5%-61.2%), 19.5% (13.8% -25.9%), and 2.0% (0.45%–5.9%), respectively. This is lower than the overall 1 – and 2 year survival rates reported in Taiwan, which were 40.3% and 22.9%, respectively) (7). The 5-year overall survival rate was 45.7 ± 0.7% in a Korean study, which is much higher than the 2-year survival rate in our study. This could be due to a delay in diagnosis and lack of standard treatments in our setup (20). Only 17 (7.4%) patients were alive at the time of analysis. The status of 43 (18.7%) patients was unknown. Of the 170 patients who died, 166 (72.2%) died at home, while the remaining four (1.7%) died in the hospital.

Consistent with other studies, survival time varied among different categories of covariates such as histologic subtypes, pathologic and clinical stage at presentation, and treatment with surgery or chemotherapy (27,28). The median survival times for stages I, II, III, IVA, and IVB were 8, 4 (95%CI:2-5.7), 9 (6.6 -11), 6 (4.8-7), and 4 (2.6–5.4) respectively. Patients with no distant metastases (IVA) during diagnosis had better survival than those with distant metastases (log-rank test, P = 0.0158). Treatment with chemotherapy and/or radiotherapy was associated with improved survival in Kaplan-Meier analysis (P= 0.032 and 0.041, respectively). This finding is consistent with the results of several other studies. Consistent with other studies, SCC is associated with a poorer prognosis (27). Bone and distant lymph node metastases have been reported to be associated with poor prognosis (29). In our study, we found brain metastases resulting in poor outcome.

### Strength

The present study has several strengths. We used a prospective cohort study design that enabled us to obtain full information on the variables and follow up patients to detect event occurrences. Second, the referral hospital in which this study was conducted was the only referral hospital in Ethiopia with available radiotherapy treatment. This presumably provides a good representation of the country.

### Limitation

Cancer-specific death was not assessed because almost all patients died at home, with no record of the immediate cause of death. As a result, deaths due to esophageal cancer might have been overestimated, leading to outcome ascertainment bias. Nevertheless, because misclassification is independent of prognostic factors, the effect on hazard ratios is negligible.

### Conclusion

In Ethiopia, patients with esophageal cancer do not receive multimodal treatment. This resulted in very low six month, one-, and two-year survival. Despite a very low overall survival, patients who received chemotherapy, radiotherapy, or surgery showed better survival than those who did not receive any treatment. We recommend improving the survival of patients with esophageal cancer through timely initiation of multimodal treatment options. Dysphagia is a major symptom that requires immediate palliation and improves quality of life before other interventions. We recommend expanding the service to include endoscopic interventions with metallic or plastic tubes (stents).

### Generalizability

The study results are generally applicable to other settings, as the sample size is large and the study was conducted at a cancer referral hospital with radiotherapy services available. The prospective cohort design also enabled us to collect data closely and accurately, making the results more representative and generalizable. However, the results may not be applicable in settings where the multimodality treatment was available for patients specifically in developed nations. Additionally, the study results may be affected by the presence of confounding factors that were not accounted for in the analysis, such as social and economic status.

## Data Availability

The data used for this research will be available upon request.

## Acknowledgments

We wish to express our sincere gratitude to our medical chart room workers, nurses, residents, and physicians for providing great support during the research process despite their busy schedule. Special thanks go to Mrs. Fikirte, Miss. Muluwork, Mr. Nemie, and Mrs. Ayalnesh for their invaluable help.

## Funding

This study was funded by the Addis Ababa University School of Public Health through the esophageal cancer thematic research project award with support from Martin-Luther-University Halle-Wittenberg, Halle (Saale), Germany. The principal investigator, Jilcha Diribi Feyisa, received the fund for this project. The funders had no role in study design, data collection and analysis, decision to publish, or preparation of the manuscript.

## Supplemental

1. Supplemental Fig - Kaplan Meier plot of survival experience of esophageal cancer patients based on stages (A) and presence of metastases (B) at a presentation, Tikur Anbessa Specialized Hospital, Addis Ababa, Ethiopia 2018 – 2020.
2. Supplemental Table 1: Stage at the presentation of esophageal cancer patients at Tikur Anbessa Specialized Hospital, Addis Ababa, Ethiopia 2018 – 2020.
3. Supplemental Table 2: Type of treatment given for esophageal cancer patients at Tikur Anbessa Specialized Hospital, Addis Ababa, Ethiopia 2018 – 2020.
4. Supplemental Table 3: Tabulated presentation of bivariate Cox regression analysis of esophageal cancer patients at Tikur Anbessa Specialized Hospital, Ethiopia.
5. Supplemental Table 4: Tabulated presentation of multicollinearity test of variables having gross association on bivariate Cox regression for esophageal cancer patients in TASH, Ethiopia.
6. Supplemental Table 5: Variables with an association to survival on multivariate Cox regression of esophageal cancer patients at Tikur Anbessa Specialized Hospital, Addis Ababa, Ethiopia 2018 – 2020.

## Notes

### Competing Interest Statement

The authors have declared no competing interest.

### Author Declarations

Ethical clearance was obtained from the Addis Ababa University Institutional Review Board (IRB) (IRB Reference Number: 096/17/SPH). Permission to review patient charts and contact patients during their visits was obtained from the oncology department. Written informed consent to participate in the study was obtained during the hospital visits. Confidentiality of the information was maintained throughout the study by excluding names as identification in the data extraction form and the data used only for the purpose of the conducted study.

